# Prodromal symptoms of rheumatoid arthritis in a primary care database: variation by ethnicity and socioeconomic status

**DOI:** 10.1101/2023.11.13.23298446

**Authors:** Alexander d’Elia, Aliaksandra Baranskaya, Shamil Haroon, Ben Hammond, Nicola J Adderley, Krishnarajah Nirantharakumar, Joht Singh Chandan, Marie Falahee, Karim Raza

## Abstract

**Objectives:** To assess whether prodromal symptoms of rheumatoid arthritis (RA), as recorded in the Clinical Practice Research Datalink Aurum (CPRD) database of English primary care records, differ by ethnicity and socioeconomic status.

**Methods:** A cross-sectional study to determine the coding of common symptoms (≥0.1 % in the sample) in the 24 months preceding RA diagnosis in CPRD Aurum, recorded between January 1^st^ 2004 to May 1^st^ 2022. Eligible cases were adults with a code for RA diagnosis. For each symptom, a logistic regression was performed with the symptom as dependent variable, and ethnicity and socioeconomic status as independent variables. Results were adjusted for sex, age, BMI, and smoking status. White ethnicity and the highest socioeconomic quintile were comparators.

**Results:** In total, 70115 cases were eligible for inclusion, of which 66.4 % female. Twenty-one symptoms were coded in more than 0.1 % of cases so were included in the analysis. Patients of South Asian ethnicity had higher frequency of codes for several symptoms, with the largest difference by odds ratio being muscle cramps (OR 1.71, 1.44-2.57) and shoulder pain (1.44, 1.25-1.66). Patients of Black ethnicity had higher prevalence of several codes including unintended weight loss (2.02, 1.25-3.28) and ankle pain (1.51, 1.02-2.23). Low socioeconomic status was associated with morning stiffness (1.74, 1.08-2.80) and falls (1.37, 2.03-1.82)

**Conclusion:** There are significant differences in coded symptoms between demographic groups, which must be considered in clinical practice in diverse populations and to avoid algorithmic bias in prediction tools derived from routinely collected healthcare data.

**Key messages:**

- There are differences in symptom reporting in new onset rheumatoid arthritis across ethnic groups.
- These differences should be considered in clinical practice in diverse populations.
- The findings are relevant in avoiding bias in prediction tools derived from healthcare data.

## Introduction

Rheumatoid arthritis (RA) is a common immune-mediated inflammatory condition with an adult prevalence of 0.8 % in the UK (1). Patients typically present in primary care before being referred to and diagnosed by rheumatologists (2). Treatment within three months of symptom onset is associated with improved clinical outcomes, including higher chances of sustained remission, reduced joint destruction, and reduction of extra-articular disease manifestations (3). Despite this, a recent UK audit found that half of all patients experienced symptoms for longer than six months prior to referral (as reported by secondary care clinicians) (2). Similarly, an older study found that a quarter of patients experience symptoms for more than 66 weeks before seeing a rheumatologist (4) An increase in primary care consultations in the two years preceding a diagnosis of RA has been reported (5) and even after being seen in primary care, 44% of patients are still not referred within the target of three working days (2), and a Danish study of RA patients found that 25% of RA patients had five or more GP consultations before RA was considered as a cause for their symptoms (6). The above suggest scope for earlier identification and referral of suspect cases to secondary care.

Meanwhile, there are well-documented ethnic and socio-economic disparities in clinical outcomes for RA (7-9), suggesting a lack of health equity along the patient pathway. There is evidence that ethnicity and socio-economic status influence the symptomatic presentation to primary care (8, 10) and patients of non-White ethnicity and low socioeconomic status may be more likely to present with “atypical” musculoskeletal symptoms than their White or more affluent counterparts. Such presentations of prodromal RA may pose a diagnostic challenge, contributing to referral lag (2). This may be further compounded by multimorbidity, which is associated with both ethnicity and socioeconomic status, and makes recognition of early RA more difficult, as new RA-related symptoms may be incorrectly attributed to pre-existing conditions (11).

Improved understanding of how the symptomatology of early RA varies with ethnicity and socioeconomic status is needed to address diagnostic delay, and ultimately reduce health inequities. Development of data-driven clinical prediction models could contribute to earlier referral, diagnosis and treatment (12). However, under-representation of subpopulations within the datasets used to build such prediction models, in combination with demographic differences in presentation, may result in less accurate predictions for some groups. For example, Chen et al. discussed the potential implication of such imbalance in relation to intensive-care-mortality prediction, which was shown to be more accurate for White men compared to women and patients of minority ethnicities (13). This algorithmic bias (14, 15) may further contribute to diagnostic delay and worsen health inequities. As the present study utilises the large Clinical Practice Research Datalink Aurum (CPRD Aurum) dataset (16), our findings offer insight into the risk of algorithmic bias in RA-prediction models built on the same dataset.

## Aim

To assess whether the prodromal symptoms of rheumatoid arthritis (RA), as recorded in English primary care records in the CPRD Aurum database, differ by ethnicity and socioeconomic status. The analysis aims to offer insight into demographic differences in early RA presentations, and to highlight the risk of algorithmic bias in tools developed from CPRD Aurum data.

## Methods

We conducted a cross-sectional study in the CPRD Aurum database investigating variations in the frequency of common (prevalence ≥0.1 %) symptoms coded in the 24 months preceding a recorded RA diagnosis. Variations were subsequently investigated by ethnicity and socioeconomic status.

CPRD Aurum is an anonymised database of observational clinical routine data (OCRD). It consists of primary care medical records of over 13 million actively registered patients in general practices in England and Northern Ireland that use the EMIS clinical information system. It captures data on patient demographics, diagnoses, symptoms, prescriptions, referrals and laboratory results. Structured data on diagnoses, symptoms and referrals are recorded using SNOMED CT coding terminology. Data are released regularly for research purposes, and this study utilised data from the May 2022 release (16).

Socioeconomic status was defined by the English Indices of Multiple Deprivation (IMD). IMD is a composite measure to quantify socioeconomic deprivation and consists of measures of income, employment, health, crime, barriers to housing and services, and living environment. All in turn are made up of several indicators. IMD data do not represent individuals but rather localities which in this study was a Lower Layer Super Output Area (LSOA) encompassing on average 1500 persons. The IMD data used are provided by CPRD, with IMD quintiles assigned to each individual based on LSOA of residence from the 2019 (latest as of June 2023) release of IMD (17). Quintile 1 represents patients living in the 20% most deprived localities.

The study period covered incident cases of RA registered from 1 January 2004 until 1 May 2022 (from the start of CPRD Aurum data until the working copy was extracted). RA was defined using existing code lists from previous work in CPRD Aurum (5). The following inclusion criteria were applied: adults (≥18 years) registered at practices in England with linked IMD data (not available for Northern Ireland), documented ethnicity, incident diagnosis of RA during the study period and at least 24 months registration time at the respective practice before the date of RA diagnosis. The duration of the prodromal phase was set to 24 months based on consultation with local rheumatology experts and previous research that showed that a large proportion of patients experience symptoms for >12 months prior to diagnosis (18).

### Exposures

The exposures were ethnicity and IMD quintile. Ethnicity categories were defined by the five high-level groups recorded in the CPRD Aurum dataset: White, South Asian, Black, Mixed and Other.

### Symptoms and code lists

The symptoms included were initially derived from a CPRD Aurum-based descriptive study by Muller et al (2019) (19) on the prevalence of prodromal symptoms of RA. This was further expanded by an exploratory review of prodromal RA symptoms (Supplementary material 1). These searches resulted in a list of 36 prodromal symptoms (Supplementary material 2). Where available, existing CPRD Aurum SNOMED CT code lists generated from prior work by the research team were utilised to capture symptom occurrence. For joint related symptoms, the broad categories used by Muller et al, such as “hand problems”, were subdivided into the cardinal features of rheumatoid arthritis: pain, stiffness and swelling. New code lists were developed for these symptoms according to the following principles:

- Anatomical consideration: e.g., for “hand pain”, all joint areas of hand were included.
- Biological plausibility: e.g. “jaw pain” is a known prodromal symptom, but “jaw swelling” is not and was not included. “Foot swelling” was excluded due to inability to distinguish synovial swelling (which may relate to RA) from the common and unspecific foot oedema.
- Code exclusivity: codes were mutually exclusive in code lists. This was checked when code lists were provisionally completed, and in cases of duplication, a joint decision was made on inclusion, as per the above criteria.

Example code lists can be found in Supplementary material 3 and the complete set is available on request.

Symptoms coded in more than 0.1 % of cases within the 24 months preceding the diagnosis of RA (equivalent to n≥70 occurrences) were included in the analysis.

### Co-variates

Sex, age, body mass index (BMI) and smoking status were included in the model as co-variates. Sex was treated as binary as per the data in CPRD Aurum. Age was included as four groups (18-30 years, 31-50 years, 51-70 years and >70 years). Sex and, in particular, age bring significant physiological differences which may explain symptom variation. Sex- and age-differences in symptomatology are already well reported (20). BMI was included as it is known to affect musculoskeletal symptoms (21) and varies with ethnicity and socioeconomic status (22). BMI was analysed categorically as per the following groups: <18.5kg/m2 (underweight), 18.5-24.9kg/m2 (normal weight), 25-29.9kg/m2 (overweight), 30-39.9kg/m2 (obese) and ≥40kg/m2 (morbidly obese). Smoking is also known to correlate with both symptoms and prevalence of RA (23), and was included in the regression models as: current smoker, ex-smoker and never smoked. Smoking status was ascertained from CPRD Aurum data using the method from Subramanian et al (2022) (24).

### Statistical methods

Data were extracted from CPRD Aurum using DExtER, an automated epidemiology software platform developed at the University of Birmingham (25). Statistical analysis was then performed in Stata version 14 (26). For each case (i.e. patient), all included symptoms were given a duration variable denoting the time span from the recording of the symptom and the diagnosis date, and only symptoms occurring ≤24 months before diagnosis were included. A binary logistic regression was conducted for each of the 21 included symptoms, including the exposures and covariates as independent variables and the given symptom as the dependent variable.

Results were reported as the odds ratios (OR) of the comparative prevalence of symptoms preceding diagnosis in a subset of the population, grouped by ethnicity and IMD quintile compared to the prevalence in the largest ethnicity (White) and to the least deprived IMD quintile 5. This was adjusted for the confounders of sex, age group, BMI category and smoking status. The risk of type-1 error due to multiple regression models was addressed by incorporating a Bonferroni correction to adjust the *p* value thresholds for statistical significance (21 regression models gave *p*<0.0024 for 95 % confidence), and subsequently results are expressed with 99.76 % confidence interval.

Three supplementary analyses were conducted: a) comparison of the studied symptoms with a matched non-RA population, to assess whether differences in symptomatology reflect differences in RA presentation or other differences between ethnic groups which are unrelated to RA; b) comparison of adjusted and non-adjusted odds ratios for “any symptom” to assess the impact of the confounders; and c) stratified analyses for “any symptom” by ethnicity and IMD quintile to assess the interaction between ethnicity and IMD (for further detail see Supplementary material 4).

### Missing data

Cases with missing ethnicity and IMD were excluded as these datapoints were central to the aim. Missing data on BMI category and smoking status were replaced by a “missing” value and included. Implausible BMI (<10, >100 kg/m2) was treated as missing.

### Patient and public involvement

A panel of five patient research partners contributed to the development of the grant application that partially funded this research. Development of the current research objectives and interpretation of findings was supported by monthly project meetings, in which a patient research partner participated. This manuscript was reviewed, proofread, and approved by a patient research partner.

### Ethics

This study and the use of CPRD Aurum and linked IMD data was approved by the CPRD Research Data Governance board, reference number 22_002367. The study was conducted in accordance with the recommendations for physicians involved in research on human subjects adopted by the 18th World Medical Assembly, Helsinki 1964, and later revisions.

## Results

The initial dataset included 83657 cases. After excluding cases with missing data on ethnicity (n=12336) and IMD (n=1206), 70115 cases were included in the analysis.

The demographics of the study population are described in Table 1. The majority (66.4 %) of cases were female and the largest age group was 51-70 years (47.3 %), with a mean age of 60.1 years. The most common BMI group was “Overweight” (32.7 %), and the mean BMI was 25.1 (SD 4.1). Current smoking was recorded in 26.9 % of cases. The most common ethnic group was White, with 88.4 % the sample.

**Table 1:** Demographic properties of included patients. Demographic properties of included cases, and reasons for exclusion.

| **Characteristic** | **n** | **%** |
| --- | --- | --- |
| **All included** | **70115** | **100 %** |
| **Age** (mean 60.1 years, SD 14.8) |  |  |
| 18-30 | 2016 | 2.9 % |
| 31-50 | 15842 | 22.6 % |
| 51-70 | 33137 | 47.3 % |
| >70 | 19120 | 27.3 % |
| **Sex** |  |  |
| Female | 46563 | 66.4 % |
| **Ethnicity** |  |  |
| White | 62215 | 88.7 % |
| Asian | 4892 | 7.0 % |
| Black | 1965 | 2.8 % |
| Mixed | 450 | 0.6 % |
| Other | 593 | 0.9 % |
| **IMD Quintile** |  |  |
| IMD Quintile 5 (least deprived) | 13614 | 19.4 % |
| IMD Quintile 4 | 14412 | 20.6 % |
| IMD Quintile 3 | 13565 | 19.4 % |
| IMD Quintile 2 | 14063 | 20.1 % |
| IMD Quintile 1 (most deprived) | 14461 | 20.6 % |
| **Body Mass Index** (mean 25.1, SD 4.1) |  |  |
| Underweight (10-18.5) | 1221 | 1.7 % |
| Normal (18.5-25) | 19931 | 28.4 % |
| Overweight (25-30) | 22948 | 32.7 % |
| Obese (30-40) | 17494 | 25.0 % |
| Morbidly obese (>40) | 2907 | 4.2 % |
| (Missing data) | 5614 | 8.0 % |
| **Smoking status** |  |  |
| Current | 18888 | 26.9 % |
| Ex-smoker | 28764 | 41.0 % |
| Never | 21171 | 30.2 % |
| (Missing data) | 1292 | 1.8 % |
| **Exclusions** |  | Of total |
| No ethnicity recorded | 12336 | 14.7 % |
| No IMD recorded | 1206 | 1.4 % |
| All | 13542 | 16.2 % |

SD = Standard Deviation. IMD = Indices of Multiple Deprivation.

Of the initial list of 36 symptoms, 21 symptoms had a prevalence ≥0.1 % (equal to ≥70 cases) and were included in the analysis (Table 2). Of the sample, 49.6 % (n=34799) of cases had one or more of the 21 eligible symptoms coded. The average number of coded symptoms per case was 0.80 (SD 1.03), ranging from 0 to 12 symptoms. After adjusting for confounders the odds ratio (OR) for having any symptom coded was higher in cases of Black (OR 1.17, 99.76 % confidence interval 1.04-1.32) and South Asian ethnicity (OR 1.16, 1.07-1.26), compared to White ethnicity. There were no significant differences by IMD quintile for prevalence of “any symptom”.

**Table 2:**
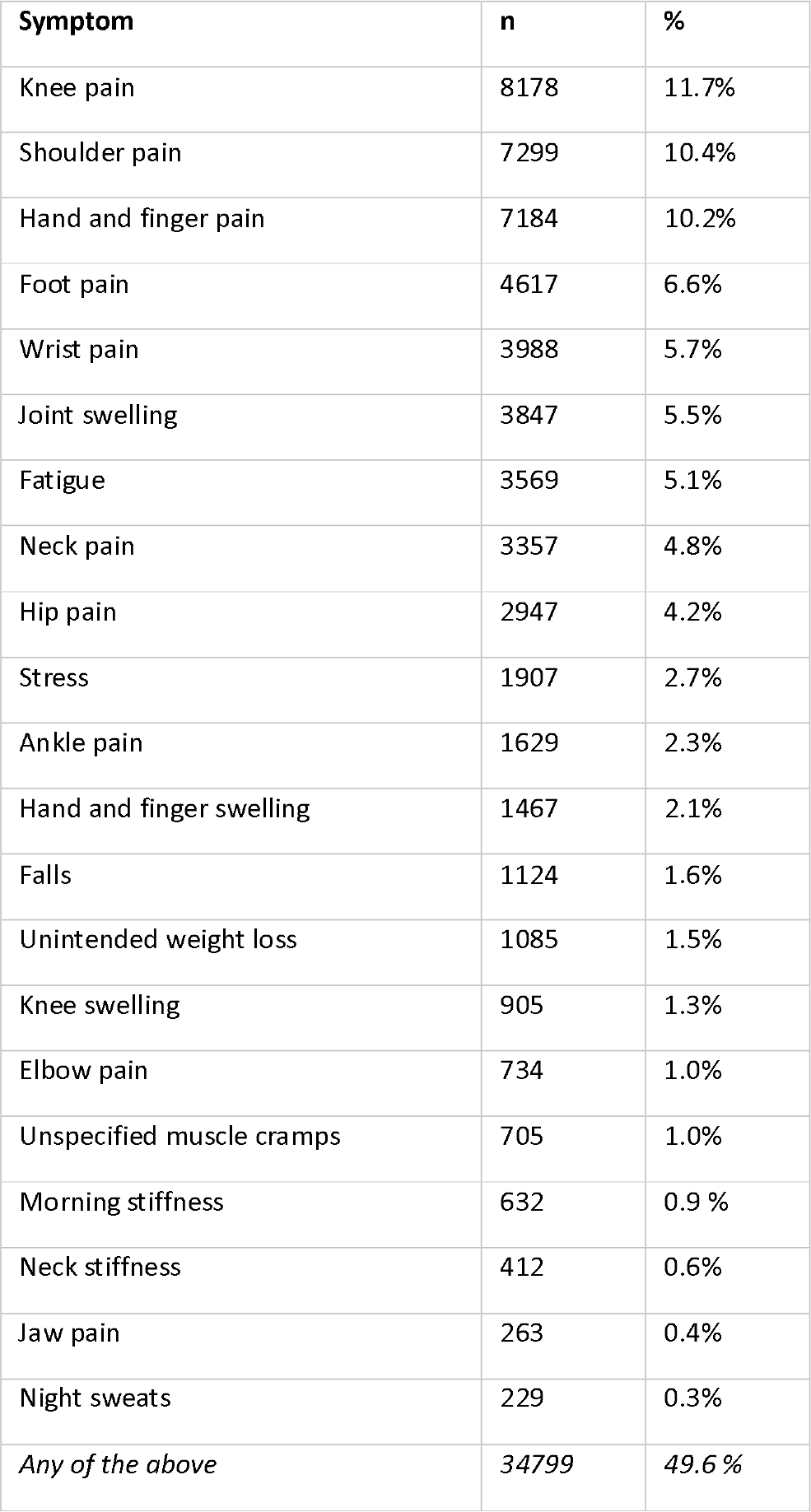
List of included symptoms. The 21 symptoms studied and their prevalence in the 24 months preceding a diagnosis of rheumatoid arthritis. Selected from the 36 initial symptoms: Only those coded in >0.1 % of the cases were included.

Statistically significant differences were found for the coding of twelve symptoms (Table 3). Cases of South Asian and Black ethnicity were more likely to have codes for knee pain (OR 1.29, 1.06-1.58 and 1.37, 1.20-1.57 respectively) and shoulder pain (OR 1.33, 1.07-1.65 and 1.44, 1.25-1.66 respectively). South Asian cases more frequently had codes for neck pain (OR 1.28, 1.04-1.57), fatigue (OR 1.28, 1.06-1.55), unspecified muscle cramps (OR 1.71, 1.14-2.57) and hand and finger pain (OR 1.16, 1.00-1.35) than any other ethnic group. However, hip pain was statistically less likely to be coded in cases of South Asian ethnicity (OR 0.66, 0.50-0.89). Ankle pain (OR 1.51, 1.02-2.23) and unintended weight loss (OR 2.02, 1.25-3.28) were more frequently coded in cases of Black ethnicity. Reporting of falls was statistically higher by the “Other” ethnicity and IMD quintile 1 (most deprived) (OR 2.14, 1.02-4.50, and 1.37, 1.03-1.82 respectively). Morning stiffness was also more frequently coded in IMD quintile 1 (OR 1.74, 1.08-2.80). Finally, jaw pain was more frequently coded in cases of Other ethnicity (OR 3.30, 1.02-10.73). See Supplementary material 5 for full results of the regression models.

**Table 3:** Symptoms with significant differences in-between groups. Overview of symptoms where a statistically significant difference was found for ethnicity (compared to White, the largest group) and IMD quintile (compared to quintile 5, least deprived). Statistical significance for p=0.0024 (p=0.05 divided by the 21 different analyses) gives a confidence interval of 99.76 % for the individual analyses.

| <u>Group</u> | <u>Symptom</u> | <u>OR</u> | <u>99.76 % CI</u> |
| --- | --- | --- | --- |
| **Black** | Unintended weight loss | 2.02 | 1.25-3.28 |
|  | Ankle pain | 1.51 | 1.02-2.23 |
|  | Shoulder pain | 1.44 | 1.25-1.66 |
|  | Knee pain | 1.37 | 1.20-1.57 |
| **South Asian** | Muscle cramps | 1.71 | 1.14-2.57 |
|  | Shoulder pain | 1.33 | 1.07-1.65 |
|  | Knee pain | 1.29 | 1.06-1.58 |
|  | Fatigue | 1.28 | 1.06-1.55 |
|  | Neck pain | 1.28 | 1.04-1.57 |
|  | Hand and finger pain | 1.16 | 1.00-1.35 |
|  | Hip pain | 0.66 | 0.50-0.89 |
| **Other ethnicity** | Jaw pain | 3.30 | 1.02-10.73 |
|  | Falls | 2.14 | 1.02-4.50 |
| **IMD quintile 1**<br>**(most deprived)** | Morning stiffness | 1.74 | 1.08-2.80 |
|  | Falls | 1.37 | 1.03-1.82 |

OR = Odds Ratio. CI = Confidence Interval.

The supplementary analyses found that: a) In an age-, sex- and medical-practice-matched control population there were similar differences in coded symptoms between ethnic groups in the non-RA control population. However, the overall symptom prevalence was much lower at 24.1 % (all ethnic groups) in the control group compared to the RA study population at 48.9 %, suggesting that the differences in coding found in the study can be attributed not only to differences in baseline symptoms, but differences in prodromal RA symptom codes. b) After excluding potential confounders from the analysis, the results were largely unaltered; thus, the included confounders had very limited impact on the results. c) The relationship between ethnic group and IMD quintile and the odds ratios for coding of “any symptom” was preserved after stratification, indicating that the results of the main analysis are unlikely to be affected by interaction between ethnicity and IMD. The results for the supplementary analyses are available in Supplementary material 4.

## Discussion

Significant differences in symptomatology (as coded) were found across twelve prodromal symptoms of RA, with higher prevalence of coded symptoms mainly in cases of South Asian and Black ethnicity. Our findings also suggest that patients of Non-white ethnicity are more likely to report general musculoskeletal symptoms (such muscle cramps and fatigue, or pain in large joints). This is clinically significant as patients presenting with non-cardinal prodromal symptoms of RA are more likely to experience longer secondary care referral delays and are thus consequently less likely to initiate treatment within 3 months of onset (27). It has previously been reported that ethnic minorities and socioeconomically disadvantaged subpopulations experience a worse functional status and impact on quality of life from RA (9), and it is possible that delayed diagnosis and treatment is a contributory factor (8). Beyond RA, these groups experience worse overall health outcomes (for example during the COVID-19 pandemic (28)), and reducing these health inequities is a priority and statutory duty for healthcare systems (29), including the English NHS which forms the setting of this analysis (30). Improving diagnostic accuracy and reducing diagnostic delay would help combat these inequities in health.

Socioeconomic deprivation was only found to correlate with increased prevalence of morning stiffness and falls, and only in IMD quintile 1 (most deprived). As such, our data suggest that socioeconomic status impacts the reporting of prodromal symptoms of RA to a lesser degree than ethnicity. However, ethnicity is a static factor whereas patients’ socioeconomic status can change throughout lifetime and its impact is more challenging to measure and interpret. There is also a well-known correlation between ethnicity and socioeconomic status, with people of minority ethnicity more likely to be socioeconomically disadvantaged (31). However, IMD quintile was not found to strongly correlate to the prevalence of symptoms in the present study and in further stratified analysis available in Supplementary material 4, and so it is likely that the majority of the effect can be explained by ethnicity. It must be remembered, however, that IMD quintile is a proxy measure of socioeconomic deprivation as it describes areas, not individuals. The demographics of the study population are in line with preceding literature on the age and sex of incident RA cases (2). White ethnicity was over-represented in comparison to national census data (32) (88.4 % vs. 81.7 %).

Beyond informing clinical practice, the results have implications for the usage of CPRD Aurum data (and similar OCRD sources) in creating clinical prediction models. If differences in symptom patterns exist between different ethnic groups (as indicated by this study), prediction models must take this into account, otherwise the predictive performance will be inferior for the populations which are numerically smaller (e.g., ethnic minorities).

Further research is required on this topic to help effectively mitigate this risk of bias in prediction models. From a clinical perspective, further research would help build on these findings to form more equitable management guidelines to facilitate earlier diagnosis of RA across all ethnic groupings.

### Strengths and limitations

This analysis presents a pragmatic approach to assess systemic demographic differences in symptomatology as reflected in coding, providing a useful starting point for more targeted research. A strength is the analysis of the CPRD Aurum dataset, enabling inclusion of a large sample size. The analysis does however have limitations. The study relies on the accuracy of symptom data in CPRD Aurum and is dependent on how symptoms are recorded by individual general practitioners, and recording patterns of general practitioners may vary across ethnic groups. The low frequency of symptoms which are known to be associated with RA suggests under-coding of symptoms in CRPD Aurum. For example, more than half of all RA patients present with painful small joints of hands (2), but in this analysis, only 10.2 % of cases had this symptom coded. The previously mentioned CPRD study by Muller et al (2019) (19) (which draw data from the parallel CPRD system CPRD GOLD) indicate this is to be expected, with a recorded frequency of finger joint pain of 16.2 % using a wider definition, again much lower than would be expected for RA. By design, the study does not differentiate between symptoms directly related to RA and symptoms related to other morbidities. Nonetheless, for the purpose of comparing prodromal symptoms across subpopulations without inferring causality, the current analysis is appropriate: if a certain group has more symptoms, the presence of those symptoms would be likely to introduce bias to a prediction model for RA based on that data. Supplementary analysis A indicated that the baseline prevalence of symptoms was similar across the ethnic groups.

Additionally, it is possible that the dataset was not large enough to test the hypothesis in the smallest groups (e.g., the smallest ethnic group, “Mixed”). Similarly, the five ethnic groups in the CPRD data used within this study encompass vastly varied ethnic subpopulations. Additionally, through relying on primary care OCRD, subpopulations less likely to be in contact with their general practitioners are likely to have been underrepresented in the analysis, potentially introducing bias from underrepresentation in this study. Finally, 14.7 % of the initial sample did not have ethnicity recorded, which may have biased the results. Linking the dataset to hospital data (i.e. CPRD HES) may have alleviated this but this was not available within the timeframe of this analysis.

## Conclusion

In this OCRD-based cross-sectional study, we have assessed the differences in symptoms recorded in the 24 months preceding a diagnosis of RA in primary care in relation to ethnicity and socioeconomic status (defined as IMD quintile). We found significant differences in symptoms coded across ethnic groups, which must be considered in clinical practice in diverse populations as well as in data-based prediction tools derived from OCRD to avoid algorithmic bias. Improved understanding of the differences in symptomatology between groups may enable targeted efforts to reduce inequities in treatment and outcomes of RA. Finally, this study provides guidance for future research into demographic differences in RA symptoms, including the underlying causalities and the clinical implications.

## Supporting information

Supplemental material 1 Scoping review

Supplemental material 2 All symptoms

Supplemental material 3 Example codelist

Supplementary material 4 Supplementary analyses

Supplemental material 5 All regression results

Supplemental material 6 RECORD checklist

## Acknowledgements

The authors are grateful to the members of the University of Birmingham Rheumatology Research Patient Partnership (http://www.birmingham.ac.uk/r2p2) for their contribution to this research.

## Data Availability

The data underlying this article will be shared on reasonable request to the corresponding author.

