## Supplemental material 1 Scoping review for "Prodromal symptoms of rheumatoid arthritis in a primary care database: variation by ethnicity and socioeconomic status"

Supplementary material 1: Exploratory review of prodromal symptoms

An explorative literature search was conducted in August 2022 to create the baseline list of included prodromal symptoms of RA. The search included peer-reviewed, published scholarship. The below table outlines sources. Detailed references on next page.

| **Category** | **Variable** |
| --- | --- |
| Demographics | Sex (Crowson et al, 2011) |
|  | Age (Xu et al, 2021) |
|  | Family History of RA* (Kronzer et al, 2021) |
|  | BMI (Mankia et al, 2021) |
|  | Level of physical activity* (Sun et al, 2021) |
|  | Educational level (Socioeconomic status) (Xu et al, 2021) |
|  | Ethnicity (Xu et al, 2021) |
| Symptoms / signs | Joint problems (including pain and swelling): (Muller et al, 2019)   - Shoulder problems - Neck problems - Foot problems - Hand problems - Jaw problems |
|  | Musculoskeletal pathologies: (Muller et al, 2019)   - Frozen shoulder - Carpal Tunnel Syndrome - Palindromic Rheumatism |
|  | Morning stiffness (Muller et al, 2019) |
|  | Unintentional weight loss (Muller et al, 2019) |
|  | Difficulty making a fist, grip weakness* (Wouters et al, 2019) |
|  | Fatigue (Wouters et al, 2019) |
|  | Altered sensations (e.g. tingling , numbness, neuropathy)* (Stack et al, 2014) |
|  | Falls (Stack et al, 2014) |
|  | Weakness* (Stack et al, 2014) |

*Symptoms/demographics not recorded reliably in CRPD Aurum and therefore excluded from the study.

Crowson, C.S., Matteson, E.L., Myasoedova, E., Michet, C.J., Ernste, F.C., Warrington, K.J., Davis, J.M., III, Hunder, G.G., Therneau, T.M. and Gabriel, S.E. (2011), The lifetime risk of adult-onset rheumatoid arthritis and other inflammatory autoimmune rheumatic diseases. Arthritis & Rheumatism, 63: 633-639. <https://doi.org/10.1002/art.30155>

Khidir S, Wouters F, van der Helm-van Mil A, van Mulligen E. 2022. The course of fatigue during the development of Rheumatoid Arthritis and its relation with inflammation: a longitudinal study.Joint Bone Spine. https://doi.org/10.1016/j.jbspin.2022.105432.

Kronzer V, Crowson CS , Sparks JA , Myasoedova E, Davis J. (2021), Family History of Rheumatic, Autoimmune, and Nonautoimmune Diseases and Risk of Rheumatoid Arthritis. Arthritis Care Res, 73: 180-187. <https://doi.org/10.1002/acr.24115>

Lingling Sun, Jiahao Zhu, Yuxiao Ling, Shuai Mi, Yasong Li, Tianle Wang, Yingjun Li, Physical activity and the risk of rheumatoid arthritis: evidence from meta-analysis and Mendelian randomization, *International Journal of Epidemiology*, Volume 50, Issue 5, October 2021, Pages 1593–1603, <https://doi.org/10.1093/ije/dyab052>

Mankia K, Siddle H, Di Matteo A, et al. A core set of risk factors in individuals at risk of rheumatoid arthritis: a systematic literature review informing the EULAR points to consider for conducting clinical trials and observational studies in individuals at risk of rheumatoid arthritis. RMD Open 2021;7:e001768. doi: 10.1136/rmdopen-2021-001768

Muller, S., S. Hider, A. Machin, R. Stack, R. A. Hayward, K. Raza and C. Mallen (2019). "Searching for a prodrome for rheumatoid arthritis in the primary care record: A case-control study in the clinical practice research datalink." Semin Arthritis Rheum **48**(5): 815-820.

van de Stadt, L. A., B. I. Witte, W. H. Bos and D. van Schaardenburg (2013). "A prediction rule for the development of arthritis in seropositive arthralgia patients." Annals of the Rheumatic Diseases **72**(12): 1920-1926.

Nikolet K den Hollander, Marloes Verstappen, Navkiran Sidhu, Elise van Mulligen, Monique Reijnierse, Annette H M van der Helm-van Mil, Hand and foot MRI in contemporary undifferentiated arthritis: in which patients is MRI valuable to detect rheumatoid arthritis early? A large prospective study, Rheumatology, 2022;, keac017, https://doi.org/10.1093/rheumatology/keac017

Stack RJ, van Tuyl LH, Sloots M, van de Stadt LA, Hoogland W, Maat B, Mallen CD, Tiwana R, Raza K, van Schaardenburg D. Symptom complexes in patients with seropositive arthralgia and in patients newly diagnosed with rheumatoid arthritis: a qualitative exploration of symptom development. Rheumatology. 2014 Sep 1;53(9):1646-53.

Wouters F, van der Giesen FJ, Matthijssen XME*, et al* Difficulties making a fist in clinically suspect arthralgia: an easy applicable phenomenon predictive for RA that is related to flexor tenosynovitis. *Annals of the Rheumatic Diseases* 2019;**78:**1438-1439.

Xia Feng, Xizhu Xu, Yanjun Shi, Xuezhen Liu, Huamin Liu, Haifeng Hou, Long Ji, Yuejin Li, Wei Wang, Youxin Wang, Dong Li, "Body Mass Index and the Risk of Rheumatoid Arthritis: An Updated Dose-Response Meta-Analysis", BioMed Research International, vol. 2019, Article ID 3579081, 12 pages, 2019. <https://doi.org/10.1155/2019/3579081>

Xu Y, Wu Q. Prevalence Trend and Disparities in Rheumatoid Arthritis among US Adults, 2005-2018. J Clin Med. 2021 Jul 26;10(15):3289. doi: 10.3390/jcm10153289. PMID: 34362073; PMCID: PMC8348893.
