## Supplemental material 2 All symptoms for "Prodromal symptoms of rheumatoid arthritis in a primary care database: variation by ethnicity and socioeconomic status"

Supplementary material 2: Included symptoms

Table of the 36 included prodromal RA symptoms, as defined by the preceding review by Muller et al (2019) (19) and modified as part of the exploratory review (see Supplementary material 1).

| **Localised musculoskeletal symptoms** | |
| --- | --- |
| Ankle issues | Jaw issues |
| *Ankle pain* | *Jaw pain* |
| *Ankle stiffness* | *Jaw stiffness* |
| *Ankle swelling* | Knee issues |
| Elbow issues | *Knee pain* |
| *Elbow pain* | *Knee stiffness* |
| *Elbow stiffness* | *Knee swelling* |
| *Elbow swelling* | Neck issues |
| Foot issues | *Neck pain* |
| *Foot pain* | *Neck stiffness* |
| *Foot stiffness* | Wrist issues |
| Hand and finger issues | *Wrist pain* |
| *Hand and finger pain* | *Wrist swelling* |
| *Hand and finger stiffness* | *Wrist stiffness* |
| *Hand and finger swelling* | Shoulder issues |
| Hip issues | *Shoulder pain* |
| *Hip pain* | *Shoulder stiffness* |
| *Hip stiffness* | *Shoulder swelling* |
| **Other musculoskeletal symptoms** | **Non-musculoskeletal symptoms** |
| *Joint swelling* | *Unintended weight loss* |
| *Unspecified muscle cramps* | *Fatigue* |
| *Morning stiffness* | *Night sweats* |
|  | *Gingivitis or periodontitis* |
|  | *Falls* |
|  | *Stress, unspecified* |
|  | *Rheumatic nodules* |
