## Supplemental material 3 Example codelist for "Prodromal symptoms of rheumatoid arthritis in a primary care database: variation by ethnicity and socioeconomic status"

Supplementary material 3: Example of code list

Example of code list to extract data from CPRD Aurum, below for symptom “Ankle Pain”. The full set of code lists is available on request from the corresponding author.

| **MEDICAL_CODE_ID** | **DESCRIPTION** | **READ_CODE** | **SNOMED_CT_CODE** | **FREQUENCY** | **DATABASE** |
| --- | --- | --- | --- | --- | --- |
| 310840017 | Ankle joint pain | N094711 | 202490009 | 65032 | CPRD_AURUM |
| 890671000006110 | Ankle/foot joint pain | N094799 | 267954009 | 16528 | CPRD_AURUM |
| 483791000006115 | Ankle pain | N245.11 | 247373008 | 1285109 | CPRD_AURUM |
| 369388017 | Ankle pain | 1M13.00 | 247373008 | 198771 | CPRD_AURUM |
| 11902321000006100 | Subtalar joint pain |  | 202491008 | 253 | CPRD_AURUM |
| 5502581000006110 | Ankle and/or foot joint pain |  | 267954009 | 20 | CPRD_AURUM |
| 310842013 | Arthralgia of subtalar joint | N094Q00 | 202491008 | 882 | CPRD_AURUM |
| 12224031000006100 | Tenderness of ankle joint |  | 299446004 | 3 | CPRD_AURUM |
| 12224071000006100 | Tenderness of subtalar joint |  | 299553005 | 1 | CPRD_AURUM |
| 8018171000006110 | Chronic ankle pain |  | 51881000119109 | 5 | CPRD_AURUM |
| 5867071000006110 | Ankle joint - painful on movement |  | 299447008 | 38 | CPRD_AURUM |
