## Supplementary material 4 Supplementary analyses for "Prodromal symptoms of rheumatoid arthritis in a primary care database: variation by ethnicity and socioeconomic status"

a) Odds ratio of any symptom in study population and matched controls

The study population is compared with a matched control group extracted from the same CRPD Aurum dataset, matched by age (±1 year), sex and primary care practice but without a diagnosis of RA. The analyses were adjusted logistic regressions with “having any of the included 21 symptoms” as the binary dependent, ethnicity as independent, and age, sex, BMI and smoking status as covariates in the same manner as the main analysis. This was conducted separately for individuals with RA and matched controls without RA. Note that socioeconomic status/IMD is not included, due to these data not being available for the control group at the time of analysis. This explains why the sample size in the exposed population is higher than the sample in the main analysis (no cases excluded due to lack of IMD data). However, the main analysis indicated that IMD had very limited impact on the OR of reporting symptoms of prodromal RA.

Coded symptoms were overrepresented in cases of Black and South Asian ethnicity to a similar degree in the control population as in the exposed cohort. However, the overall symptom prevalence was much lower at 24.1 % (all ethnic groups) in the control group compared to the RA study population at 48.9 %, suggesting that the differences in symptoms found in this study can be attributed to not only differences in baseline symptoms, but actual differences in prodromal RA symptoms.

Table: Adjusted odds ratio of having any of the 21 included symptoms in the study population and in a non-RA control group matched by age (+- 1 year), sex and primary care practice.

|  | **Ethnicity** | **OR** | **p** | **95 % CI** | |
| --- | --- | --- | --- | --- | --- |
| **Exposed** | *Black* | 1.21* | <0.01 | 1.10 | 1.32 |
| n=71320  Prevalence of any symptom:  48.9 % (n=34687) | *Mixed* | 0.95 | 0.53 | 0.81 | 1.11 |
|  | *Others* | 1.10 | 0.29 | 0.92 | 1.32 |
|  | *South Asian* | 1.18* | <0.01 | 1.11 | 1.25 |
|  | *White* | 1.00 | (base) |  |  |
| **Controls** | *Black* | 1.12* | 0.03 | 1.01 | 1.24 |
| n=67483  Prevalence of any symptom:  24.1 % (n=16257) | *Mixed* | 1.06 | 0.58 | 0.87 | 1.28 |
|  | *Others* | 0.98 | 0.87 | 0.79 | 1.22 |
|  | *South Asian* | 1.35* | <0.01 | 1.26 | 1.45 |
|  | *White* | 1.00 | (base) |  |  |

OR = Odds Ratio. CI = Confidence Interval. RA = Rheumatoid Arthritis.
* Statistically significant at p<0.05.

b) Comparison of adjusted and non-adjusted regression model

Results of regression model for odds ratio (OR) of having any of the 21 included symptoms. Adjusted model includes BMI, sex, age and smoking status. Asterix denotes statistical significance (p<0.05). The exclusion of confounders (non-adjusted analysis) did not affect the results to a significant degree.

| **Adjusted** | **OR** | **p** | **95 % CI** |  |
| --- | --- | --- | --- | --- |
| Black | 1.17 | 0.00 | 1.07 | 1.29 |
| Mixed | 1.06 | 0.55 | 0.88 | 1.28 |
| Others | 0.99 | 0.91 | 0.84 | 1.17 |
| South Asian | 1.16 | 0.00 | 1.09 | 1.23 |
| White | 1.00 | . | . | . |
| IMD q5 | 1.00 | . | . | . |
| IMD q4 | 1.03 | 0.27 | 0.98 | 1.08 |
| IMD q3 | 1.01 | 0.59 | 0.97 | 1.06 |
| IMD q2 | 0.99 | 0.63 | 0.94 | 1.04 |
| IMD q1 | 1.01 | 0.58 | 0.97 | 1.06 |
| **Unadjusted** | **OR** | **p** | **95 % CI** |  |
| Black | 1.17 | 0.00 | 1.07 | 1.28 |
| Mixed | 1.02 | 0.85 | 0.85 | 1.23 |
| Others | 0.96 | 0.65 | 0.82 | 1.13 |
| South Asian | 1.11 | 0.00 | 1.05 | 1.18 |
| White | 1.00 | . | . | . |
| IMD q5 | 1.00 | . | . | . |
| IMD q4 | 1.03 | 0.27 | 0.98 | 1.08 |
| IMD q3 | 1.02 | 0.47 | 0.97 | 1.07 |
| IMD q2 | 0.99 | 0.80 | 0.95 | 1.04 |
| IMD q1 | 1.03 | 0.25 | 0.98 | 1.08 |

OR = Odds Ratio. CI = Confidence Interval. IMDq = IMD quintile.

c) Stratified regression by ethnicity and by IMD quintile

Regression model for presence of any of the 21 included symptoms, exploring association with IMD quintile stratified by ethnicity (left) and association with ethnicity stratified by IMD quintile (right). Significance set to p<0.05. The stratification by ethnicity and IMD indicated that IMD has little impact on coded symptoms and that the larger effect lies with ethnicity.

| **Stratified by ethnicity, adjusted.** | | | | |  | **Stratified by IMD quintile (IMD q), adjusted.** | | | | |
| --- | --- | --- | --- | --- | --- | --- | --- | --- | --- | --- |
| **BLACK** | or | P | 95% CI | |  | **IMD q5 (least deprived)** | OR | p | 95% CI | |
| IMD q5 | 1 |  |  |  |  | Black | 1.614 | 0.048 | 0.864 | 3.015 |
| IMD q4 | 1.029 | 0.081 | 0.268 | 1.288 |  | Mixed | 0.715 | 0.308 | 0.306 | 1.669 |
| IMD q3 | 1.022 | 0.146 | 0.337 | 1.353 |  | Others | 1.24 | 0.388 | 0.653 | 2.355 |
| IMD q2 | 1.004 | 0.381 | 0.415 | 1.542 |  | South Asian | 1.212 | 0.03 | 0.965 | 1.521 |
| IMD q1 | 1.031 | 0.293 | 0.398 | 1.473 |  | White | 1 | . | . | . |
| **SOUTH ASIAN** |  |  |  |  |  | **IMD q4** |  |  |  |  |
| IMD q5 | 1 |  |  |  |  | Black | 0.91 | 0.608 | 0.567 | 1.461 |
| IMD q4 | 1.198 | 0.672 | 0.399 | 3.597 |  | Mixed | 0.83 | 0.474 | 0.426 | 1.619 |
| IMD q3 | 1.203 | 0.643 | 0.431 | 3.356 |  | Others | 0.978 | 0.911 | 0.589 | 1.625 |
| IMD q2 | 1.662 | 0.183 | 0.622 | 4.442 |  | South Asian | 1.262 | 0.004 | 1.024 | 1.555 |
| IMD q1 | 1.972 | 0.082 | 0.721 | 5.388 |  | White | 1 | . | . | . |
| **WHITE** |  |  |  |  |  | **IMD q3** |  |  |  |  |
| IMD q5 | 1 |  |  |  |  | Black | 1.027 | 0.824 | 0.757 | 1.393 |
| IMD q4 | 1.028 | 0.257 | 0.965 | 1.096 |  | Mixed | 0.9 | 0.604 | 0.533 | 1.52 |
| IMD q3 | 1.018 | 0.477 | 0.954 | 1.088 |  | Others | 0.973 | 0.885 | 0.603 | 1.572 |
| IMD q2 | 0.976 | 0.357 | 0.914 | 1.044 |  | South Asian | 1.142 | 0.049 | 0.96 | 1.358 |
| IMD q1 | 1.023 | 0.386 | 0.957 | 1.093 |  | White | 1 | . | . | . |
| **MIXED** |  |  |  |  |  | **IMD q2** |  |  |  |  |
| IMD q5 | 1 |  |  |  |  | Black | 0.91 | 0.608 | 0.567 | 1.461 |
| IMD q4 | 1.198 | 0.672 | 0.399 | 3.597 |  | Mixed | 0.83 | 0.474 | 0.426 | 1.619 |
| IMD q3 | 1.203 | 0.643 | 0.431 | 3.356 |  | Others | 0.978 | 0.911 | 0.589 | 1.625 |
| IMD q2 | 1.662 | 0.183 | 0.622 | 4.442 |  | South Asian | 1.262 | 0.004 | 1.024 | 1.555 |
| IMD q1 | 1.972 | 0.082 | 0.721 | 5.388 |  | White | 1 | . | . | . |
| **OTHER** |  |  |  |  |  | **IMD q1 (most deprived** |  |  |  |  |
| IMD q5 | 1 |  |  |  |  | Black | 1.614 | 0.391 | 0.864 | 3.015 |
| IMD q4 | 0.835 | 0.576 | 0.364 | 1.917 |  | Mixed | 0.715 | 0.235 | 0.306 | 1.669 |
| IMD q3 | 0.817 | 0.519 | 0.364 | 1.832 |  | Others | 1.24 | 0.309 | 0.653 | 2.355 |
| IMD q2 | 0.839 | 0.556 | 0.388 | 1.811 |  | South Asian | 1.212 | 0.107 | 0.965 | 1.521 |
| IMD q1 | 0.802 | 0.474 | 0.362 | 1.775 |  | White | 1 | . | . | . |

IMD q = Indices of Multiple Deprivation, quintile. CI = Confidence Interval.
